# A Counterfactual Graphical Model Reveals Economic and Sociodemographic Variables as Key Determinants of Country-Wise COVID-19 Burden

**DOI:** 10.1101/2020.06.16.20132563

**Authors:** Tavpritesh Sethi, Saurabh Kedia, Raghav Awasthi, Rakesh Lodha, Vineet Ahuja

## Abstract

**Importance:** Insights into the country-wise differences in COVID-19 burden can impact the policies being developed to control disease spread.

**Objective:** Present study evaluated the possible socio-economic and health related factors (and their temporal consistency) determining the disease burden of COVID-19.

**Design:** A retrospective analysis for identifying associations of COVID-19 burden.

**Setting:** Data on COVID-19 statistics (number of cases, tests and deaths per million) was extracted from the website https://www.worldometers.info/coronavirus/ on 10^th^ April and 12^th^ May. Variables obtained to estimate the possible determinants for COVID-19 burden included economic-gross domestic product; socio-demographic-Sustainable Development Goals, SDGs indicators related to health systems, percentage Chinese diaspora; and COVID-19 trajectory-date of first case in each country, days between first reported case and 10^th^ April, days between 100^th^ and 1000^th^ case, and *government response stringency index* (GRSI).

**Main outcomes and Measures:** COVID-19 burden was modeled using economic and socio-demographic determinants. Consistency of inferences for two time points at three levels of increasing statistical rigor using (i) Spearman correlations, (ii) Bayesian probabilistic graphical model, and (iii) counterfactual impact was evaluated.

**Results:** Countries’ economy (reflected by GDP), mainly through the testing rates, was the major and temporally consistent determinant of COVID-19 burden in the model. Reproduction number of COVID-19 was lower where mortality due to water, sanitation, and hygiene (WaSH) was higher, thus strengthening the hygiene hypothesis. There was no association between vaccination status or tuberculosis incidence and COVID burden, refuting the claims over BCG vaccination as a possible factor against COVID-19 trajectory.

**Conclusion and Relevance:** Countries’ economy, through testing power, was the major determinant of COVID-19 burden. There was weak evidence for hygiene hypothesis as a protective factor against COVID-19.

## Introduction

The novel coronavirus SARS-CoV-2, which originated in China 6 months back[1], has dramatically enveloped the global population crossing all boundaries and borders, infecting more than 5 million people and causing more than 300,000 deaths as on 21 May 2020[2]. The distinct difference in the disease burden (including infectivity and mortality) between the regions across the globe is an enigma. Despite harboring 60% of the global population, Asia accounts for only ∼18% global cases and <10% global mortality due to COVID-19^2^. Western Europe (Italy, France, Spain, United Kingdom) and USA account for about 50% and 70% of global cases and mortality despite the fact that China continued to contribute to >80% of global cases till the end of March first week. Currently, the two most populous countries in the World-India and China (accounting for 35% global population) together account for less than 4% and 3% global cases and mortality, respectively. These observations have displayed a temporal consistency with almost similar country-wise distribution of cases over the 1-month period, highlighting the impact of consistent factors which govern these epidemiologic associations.

Several extrinsic and intrinsic factors such as international travel[3], *government response stringency index*,[4],[5] economy, hygiene, sanitation, infections, and immunization coverage status can contribute to this discrepancy. Initial reports suggested a negative correlation between BCG vaccination coverage and COVID mortality and infection rates[6],[7], however, these observations were negated subsequently due to significant bias from many confounders such as demographics, testing rates and the stage of the pandemic in each country[8],[9],[10],[11]. Several other studies have shown inconsistent or weak correlations and have not sufficiently addressed interactions or confounding[5],[12],[13],[14]. Therefore, an integrated approach to decipher the network of determinants of COVID-19 burden is needed. In this study, we discovered and quantified the global determinants of COVID-19 burden through an explainable Bayesian probabilistic graphical model (PGM) with counterfactual impact assessment.

## Methods

This retrospective analysis combined country-level COVID-19 statistics at two time points, i.e., 10^th^ April and 12^th^ May in order to check temporal consistency of associations with health indicators listed under Sustainable Development Goals (SDG). Country-level disease burden due to COVID-19 (number of cases/ million population) was the outcome variable of interest. Data on COVID-19 statistics was extracted from the website-https://www.worldometers.info/coronavirus, on the afternoon of 10^th^ April (3.50 PM GMT) and 12^th^ May[2]. Following variables were obtained-*Total cases/1 Million Population, Deaths/1 Million Population* and *Tests /1 Million Population*. The ratio of number of cases to number of tests was also calculated to control for the effect of testing on the number of cases. Trajectory indicators-*date of first case, days between first reported case and 10*^*th*^ *April, days between 100*^*th*^ *and 1000*^*th*^ *case* (as per 12^th^ May)[15], *Government response stringency index* (*GRSI*)[16] and country-wise R0 estimates obtained through an exponential fit model were included. *Gross domestic product* (*GDP*) *nominal per capita* and *GDP PPP per capita were* obtained from World Bank[17]. Information on percentage Chinese diaspora from different countries was obtained[18]. Sustainable Development Goals (SDGs) indicators were obtained from Global Burden of Disease data for the year 2019 scaled between 0 (worst) and 100 (best)[19].

In 71 countries where data for all variables were available, missingness of data was assessed on the merged data(Supplementary Figure 1) and a random forest based multivariate imputation[20] was carried out on the variables which passed the filter. K-means algorithm[21] was used to discretize continuous variables into three classes (low, medium, high) in order to facilitate interpretation and counterfactual impact evaluation.

Exploratory data analyses using pairwise Spearman correlations upon the merged dataset were conducted in R language and environment for statistical computing[22] and the strengths of associations were visualized as a heatmap (p-value threshold of 0.05). Since correlations may be biased by spurious relationships and confounding, conditional associations were learned through a structure learning algorithm[23] that learns a Bayesian PGM from data. Prior knowledge was incorporated by blacklisting the edges which could violate the arrow-of-time in the structure learning process. Hill climbing optimizer with Akaike Information Criterion score was used to select the best PGM that explained the data. Exact Inference using the belief propagation algorithm[23] was learned in order to quantify the strength of learned associations using the in-house developed Bayesian Artificial Intelligence pipeline and software[24],[25]. Counterfactual analysis for estimating quantile effects and confidence intervals of direct associations discovered from the PGM was carried out using quantile regression[26] with 100 weighted bootstraps using Counterfactual library in R[27]. Specifically, the impact of *Tests/1 Million Population* upon *Total cases/1 Million Population* and *Deaths/1 Million Population*, and *WaSH mortality* upon *R0* was quantified by switching from low to high ranges of respective covariates (Supplementary Table 1).

## Results

### 1. Associations of COVID-19 indices with health-related SDGs and socio-economic indicators

Economic indicators *GDP PPP per capita* and *GDP nominal per capita* showed a significant positive and temporally consistent correlation with *Cases/1 Million Population* and *Tests/1 Million Population* (10^th^ April-r=0.87 & 0.90, respectively; 12^th^ May-r=0.80 & 0.84, respectively, p<0.05 for all) (Figure 1a & b). This is in agreement with evidence on earlier spread of disease to developed countries with high international travel and mobility. There was a significant positive correlation between *Cases/1 Million Population* and *Tests/1 Million Population*, at both time points (10^th^ April-r=0.55, 12^th^ May-r=0.56, p<0.05 for both). Further, the *under 5 mortality rank* (higher rank means lower mortality) was positively and consistently correlated with *Tests/1 Million Population* ((10^th^ April-r=0.88, 12^th^ May-r=0.83; p<0.05) and *Cases/1 Million Population* (10^th^ April-r=0.73, 12^th^ May-r=0.82; p<0.05) (Figure 1a & b). Interestingly, the *GRSI* had a weak negative correlation with *Cases/1 Million Population, Deaths/1 Million Population* and *Tests/1 Million Population* in the dataset from April but did not have a correlation in the dataset from May (Figure 1b), indicating that these measures could have had a temporary effect.

**Figure 1:**
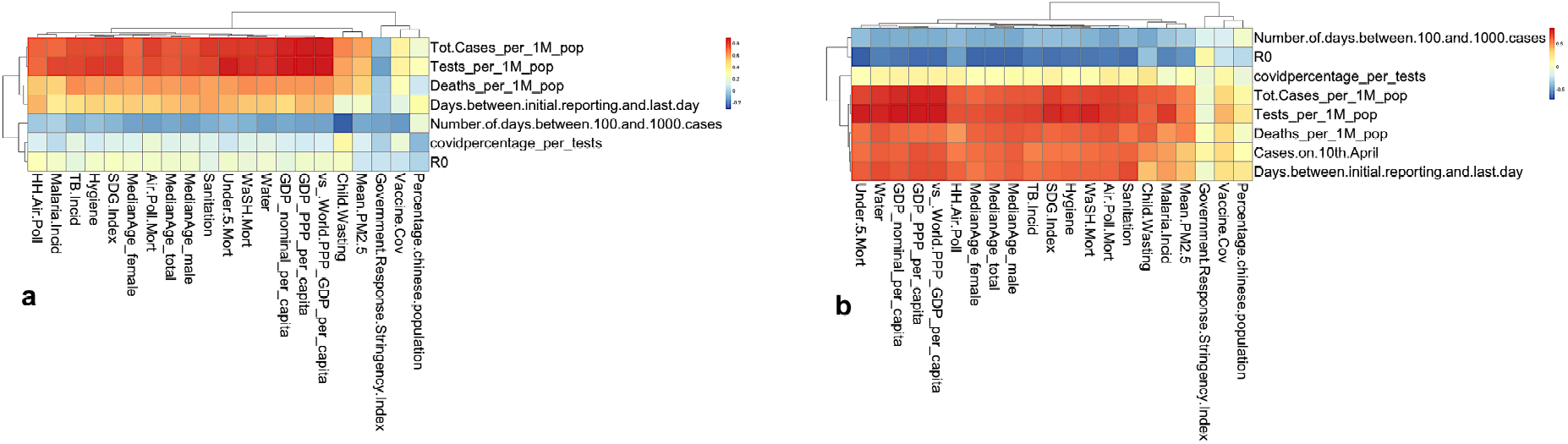
Heat map of associations of COVID-19 indicators with health-related SDGs and Socio-economic indicators on datasets analyzed on 10th April (left) and 12th May (right). At both time points, there was a consistent positive association seen between economic indicators and COVID burden. A negative association was seen between R0 and WaSH related indicators in the May dataset.

### 2. Conditional dependency structure

Structure of the Bayesian PGM learned from data represents the skeleton of direct associations and edge-directions represent asymmetric influence (Figure 2). In the model, *GDP nominal per capita* determined both *Cases/1 Million* population and *Tests/1 Million Population*. Economic features such as *GDP PPP per capita* determined *under 5 mortality rate rank* which in turn determined the state of *WaSH mortality* and *malaria incidence*. The composite indicator, *SDG index* determined *TB incidence, childhood wasting, vaccine coverage* and *sanitation* indicators and the *WaSH mortality rank* determined *R0* as seen from the flow of probabilistic influence in the learned structure.

**Figure 2.**
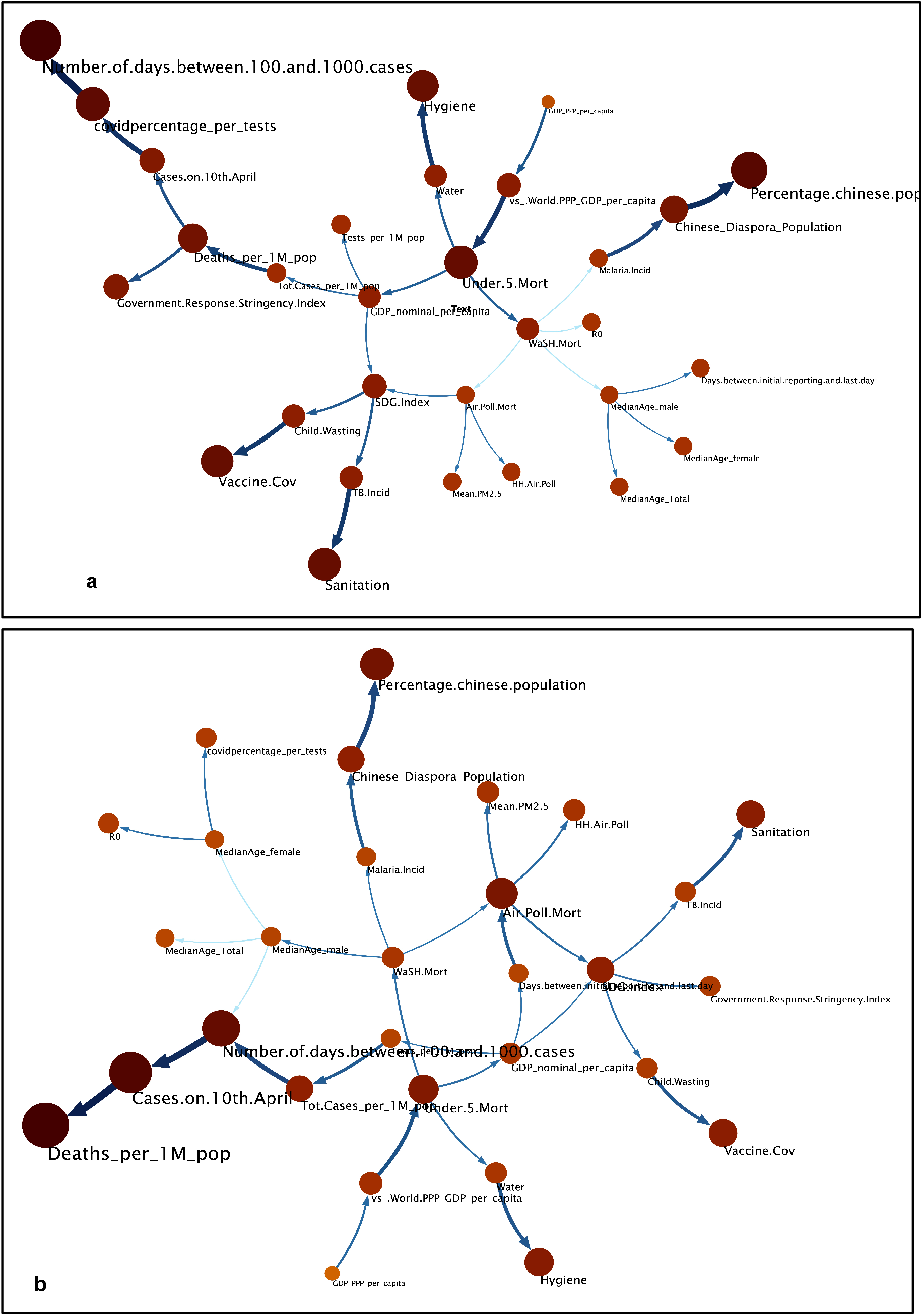
Qualitative evaluation of association structure learned as a Bayesian graphical model in the April 10^th^ (left) and May 12^th^ (right dataset). In the joint multivariate model, directed edges are consistently present between *GDP nominal per capita* and *Tests per million population*. An example of a learned relationship between *GDP nominal per capita* (red) and *Tests per million population* (green) is highlighted. Unlike pairwise associations, a Bayesian graphical model learns the joint distribution over all the variables hence avoiding spurious correlations. Some of the associations seen in Figure 1 are not seen as relationships as those are “explained away” by other associations.

### 3. Network Inference

The trend of conditional probabilities inferred from the joint probability distribution over the structure revealed a positive association between *GDP nominal per capita* and *Tests per 1Million Population* (Figure 3). This was in alignment with the findings from the pairwise correlation analysis (Figure 1) at both time snapshots. Further scenarios were simulated such as inferring GDP and SDG indicators where COVID-19 mortality was high. Figure 4 shows the log_10_ odds-ratios for COVID-19 related variables (y-axis) being in the “High” category when the covariates (x-axis) were switched from “Low” to “High”. Nearly 25 times higher likelihood of higher odds of being in the high *Total Cases/1 Million Population* was seen if the GDP was in the High segment for both the snapshots in time.

**Figure 3.**
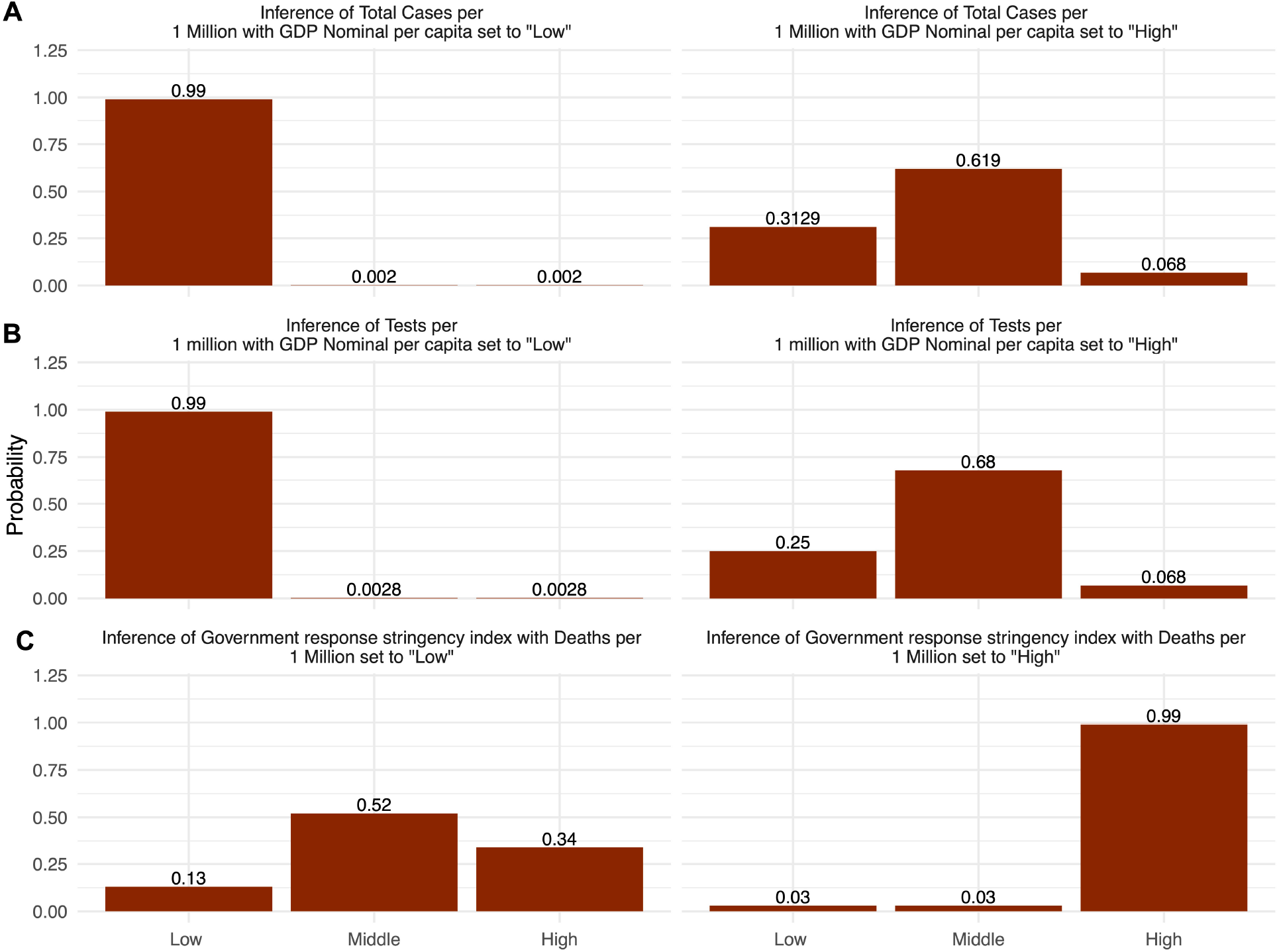
Quantitative evaluation of associations discovered in structure using Exact Inference. *GDP nominal per capita* was positively correlated with (A) *Tests/1 Million* and (B) *Total Cases/1 Million* as observed from inferred probabilities. For example, countries with low *GDP nominal per capita* are associated with low *Tests/ 1Million* (left) and vice versa (right). A positive non-monotonic correlation between *Deaths per 1Million Population* and *Government Response Stringency Index* (C) indicates that govt response followed deaths and not vice versa. However, this was a temporal response that was not observed in the May 12 analysis.

**Figure 4.**
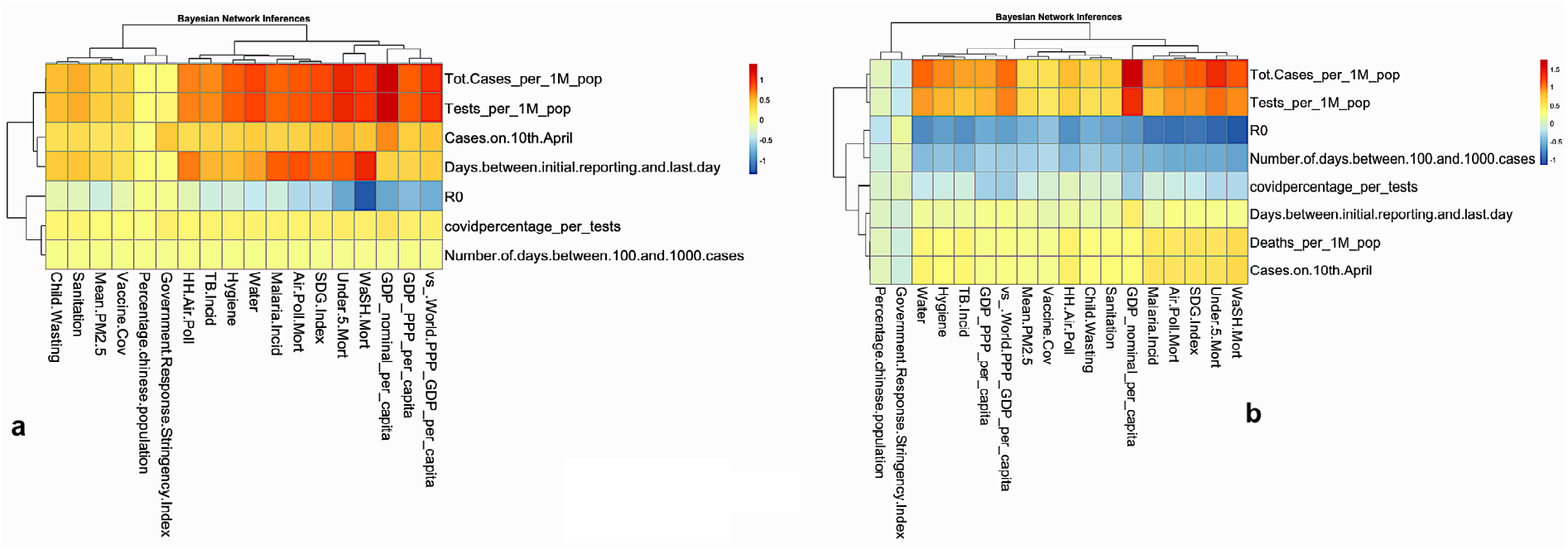
Odds-ratios for COVID-19 related variables (y-axis) being in the “High” category when the covariates (x-axis) were switched from “Low” to “High”. Up to 25 times (1.39 on log_10_ scale) higher odds of being in the high *total cases per 1 Million population* was seen if the GDP was in the High segment for both the snapshots in time. Similarly, the odds of *R0* to be high in countries with high *WaSH mortality* were up to one-tenth as compared to countries with lower *WaSH mortality* as inferred from the network.

### 4. Counterfactual Analysis

Counterfactual modeling for the rate of testing revealed that an additional 1506 (95% CI [-72, 3118]) *Cases/1 Million Population* would have been discovered if the testing rates were in the high category in the countries where it was low (Figure 5). Similarly moving a country from High to Low *covid percentage per test* would have increased the counterfactual *Number of days between 100 and 1000 cases* by up to 5.72 (95% CI [-2.08, 13.52]) days. We found that if the countries with high WaSH mortality were counterfactually switched to low WasH mortality, the R0 would have been higher by 0.67 (95% CI [-0.805, 2.15]) units. Although the CI includes zero, this weak positive effect was homogenous across the quantiles (Figure 5b). This in conjunction with the lack of association between R0 and deaths in the Bayesian PGM indicating the possible protection against spread of COVID-19 in countries with poor hygiene.

**Figure 5:**
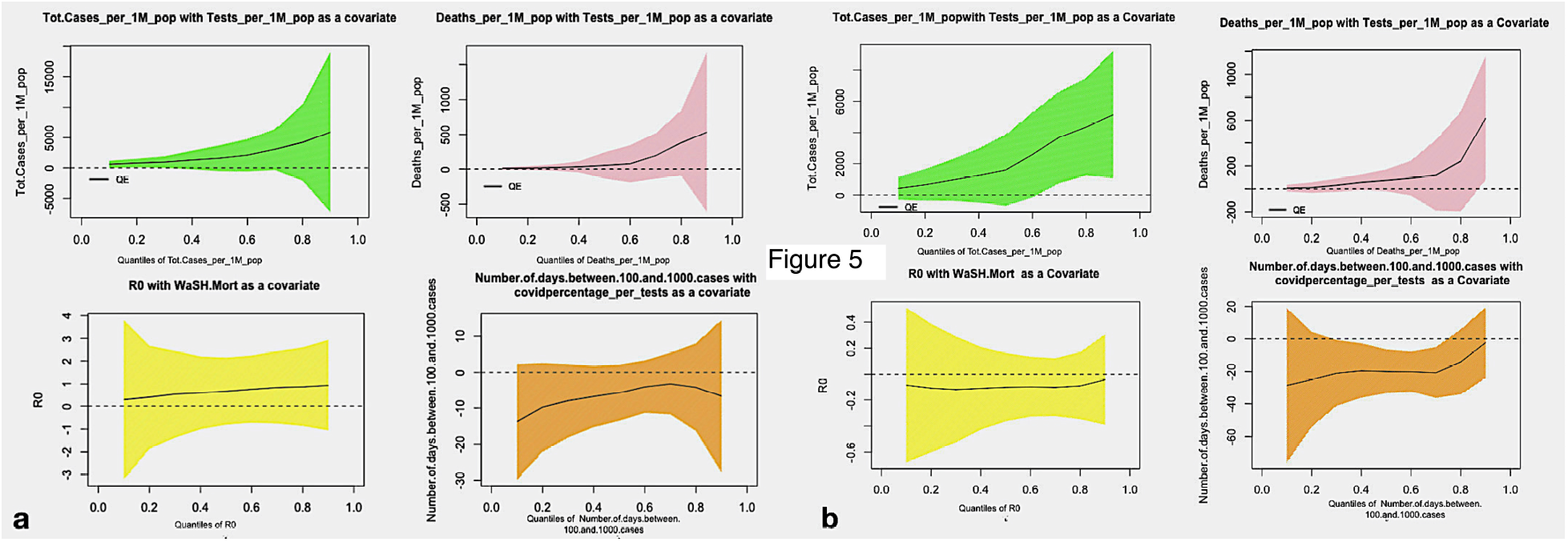
Counterfactual analysis for quantile effects and their confidence intervals calculated in 10 quantiles for a) April 10^th^ 2020, and b) May 12^th^ 2020. A consistent impact of testing rate upon increase in total cases and deaths is seen across quantile and at two time points. A similar impact of percentage COVID positive rate upon the *Number of days between between 100 and 1000* was seen whereas weak evidence for a negative impact of *WaSH mortality* upon *R0* could be observed specially in data from May 12^th^ 2020.

## Discussion

Although the COVID-19 pandemic is progressing at an exponential rate globally, the discrepant burden across different nations is intriguing clinicians, public health experts and epidemiologists. The large number of studies that have quickly accrued have failed to provide definite conclusions. Our study is an attempt to provide robust evidence for determinants of COVID-19 burden learned with a three-step pipeline from correlations, conditional dependency structure and counterfactual impact evaluation in increasing order of causal rigor. The Bayesian PGM captured several well-known influences directly from data, thus showing the abstractive power of this artificial intelligence approach. Our primary hypothesis was the dependence of COVID-19 burden on the countries’ economy, the stability of this association with time and the possible determinants of this association. Interestingly, we found an excellent correlation between *GDP nominal per capita* of a country and the COVID-19 burden at both time points (10^th^ April and 12^th^ May), 1 month apart, indicating the stability of this association even with increasing numbers and progressing pandemic. It is noteworthy that the associations and impact were stable despite the number of global cases having increased 3-fold (∼1.5 million to ∼4.5 million) and a similar increase in COVID-19 related deaths over 1-month period. The reported relationship between countries’ economy and COVID burden has been heterogeneous, with most of the results in congruence with our observation[5].

We then dissected the possible determinants of this strong association, i.e. the factors unique to both high and low-income countries which may positively or negatively affect the COVID spread. For any infectious epidemic (or pandemic), the possible determinants of disease burden include direct factors such as the susceptible population (which also has multiple determinants), and the infectivity of virus, and the indirect factors such as the rates of detection (or the number of tests performed). Although there is some evidence for mutation of virus to less virulent strains, no country-level evidence for protection against infection has been found so far[28]. Hence, the country-wide differences are expected result from a combination of testing rates and determinants of population susceptibility. As expected, we also found a strong correlation between the countries’ *GDP nominal per capita* and the *Tests/1 Million population* (again at both time points), indicating that higher income economies are testing more, which would indirectly account for higher COVID cases in these countries. The definite impact of testing on cases could also be inferred through counterfactual analysis which revealed that additional 1506 *Cases/1 Million Population* would have been discovered if the testing rates were in the high category in the countries where it was currently low. Association between testing rate and the *Cases/1 Million Population* was also significant, although not as strong as the association between GDP and the tests or *Cases/1 Million Population*. Therefore, we infer that although the economy is associated with testing and hence the number of cases, testing is not the sole determinant and there would be other possible factors guiding the strong association between economy and COVID-19 burden. Our inferences were strengthened with probabilistic machine learning, i.e. the Bayesian PGM, exact inference, and counterfactual analysis which demonstrated a positive impact of GDP on cases and tests/million. However, there was a non-monotonic correlation between tests and cases/million, with countries with a middle and high testing rate having similar number of cases (supplementary figure 2), reiterating that factors additional to testing rate are determining the disease burden, and these would be related to the susceptibility of the population.

Susceptibility of a population is likely to be determined by extrinsic factors such as international travel and the measures of social distancing and isolation, and the intrinsic factors such as the individuals’ genetic and immunological susceptibility. Although we did not evaluate the correlation between international travel and COVID-19 burden due to lack of reliable data, we believe that the international travel may have been a causal link between economy and higher rates of disease as indicated by air-travel statistics[29],[30] and collateral evidence from influenza[31]. Another extrinsic factor, GRSI, which is an indicator of the local administration’s policy on social distancing, lockdown and public closure, had a weak negative correlation with COVID cases, indicating lesser burden in countries with higher GRSI, India being a perfect example. With a GRSI of 100%, and considering the large Indian population, India had a much lower COVID burden as on the afternoon of 10^th^ April 2020. However, this relationship was not observed in the May dataset, thus indicating the temporary effect of a stringent response on the COVID burden.

Of the possible individual specific determinants, genetic and direct immunological correlations was out of scope for this paper. Hence, we looked at the indirect measures of individuals’ susceptibility to infection and disease, specifically those which are influenced by the countries’ economy, and these (sustainable Development Goals [SDGs]) were obtained from Global Burden of Disease data for the year 2019. There was a highly significant negative correlation between the under-5 mortality and tests and cases/million, indicating the higher number of cases in developed countries with lower under-5 mortality, which also had a direct association with GDP on Bayesian network analysis. Hygiene hypothesis[32], which has been correlated with higher incidence of allergic and immunologic diseases in the developed countries was also tested as a possible determinant of discrepant COVID burden. Taking WaSH mortality as a surrogate for hygiene hypothesis, we found a weak evidence for the same on counterfactual analysis, where the R0 (hence the spread) for COVID-19 increased on switching the high WaSH (poor hygiene) countries to low WaSH (better hygiene). As per the hygiene hypothesis, childhood exposure to infections resets the immune system protecting against allergic disorders, accounting for low prevalence of these diseases in developing countries. Similar immunological mechanisms could be protecting against COVID-19. BCG vaccination which trains the innate immunity against other infections[33] showed a negative association with COVID burden and deaths initially[34],[35], though these claims were refuted by subsequent studies[36]. The present study also did not find a direct association of vaccination coverage or tuberculosis incidence with COVID-19 burden, thereby negating the effect of BCG vaccination in protection against COVID-19.

Our study is strengthened by consistent association between 2 time points, 1 month apart, which is a significant time duration for a rapidly spreading disease like COVID-19. The findings inferred from the data on 10^th^ April were reiterated on 12^th^ May, validating our hypothesis and associations between possible determinants of COVID-19 burden. However, there are certain limitations. We could not calculate exact R0 due to lack of information on susceptible population and rates of recovery. We did not include other variables such as international travel, and population age, though we believe that there are other evidences supporting the associations of these variables with COVID-19 burden.

Our study provides strong evidence for the role of countries’ economy and factors such as testing rates as major determinants of COVID-19 burden. We also find that other factors in line with the hygiene hypothesis may have some role in COVID-19 burden. Finally, our approach can form the basis of further studies using a similar integrate-and-dissect analytical framework for learning granular determinants and policy to effectively mitigate the pandemic.

## Data Availability

Data generated by our research and supporting our article will be made available as soon as possible, wherever legally and ethically possible

## Author contributions

TS-Study design, Data collection, Data analysis, manuscript writing

SK-Data collection, manuscript writing

RA-Data collection, Data analysis, manuscript writing

RL and VA-Study concept, study design, manuscript writing and critical revision of the manuscript for important intellectual content

## Conflict of interest

None

## Funding

None

### Acknowledgement

This work was partly supported by the Wellcome Trust/DBT India Alliance Fellowship IA/CPHE/14/1/501504 awarded to Tavpritesh Sethi. Tavpritesh Sethi also acknowledges support from the Center for Artificial Intelligence at IIIT-Delhi.

We acknowledge Aditya Nagori for his help in preparing figures.

## Figure legends

**Low/High Ranges of Covariates in Counterfactual analysis:**

**Supplementary Table 1:** Every continuous variable has been discretized in 3 disjoint intervals for Bayesian Network analysis, we took low intervals as a low range of covariate and upper interval as high range of covariate.

| Covariates | Low Range | High Range |
| --- | --- | --- |
| Testing per 1 M population | [8, 1958) | [1958, 95718] |
| WaSH Mortality | [8.1, 36.8) | [36.8, 100] |
| Covid percentage per test | [0.11, 13.2) | [13.2, 93.3] |

**Supplementary Figure 1.**
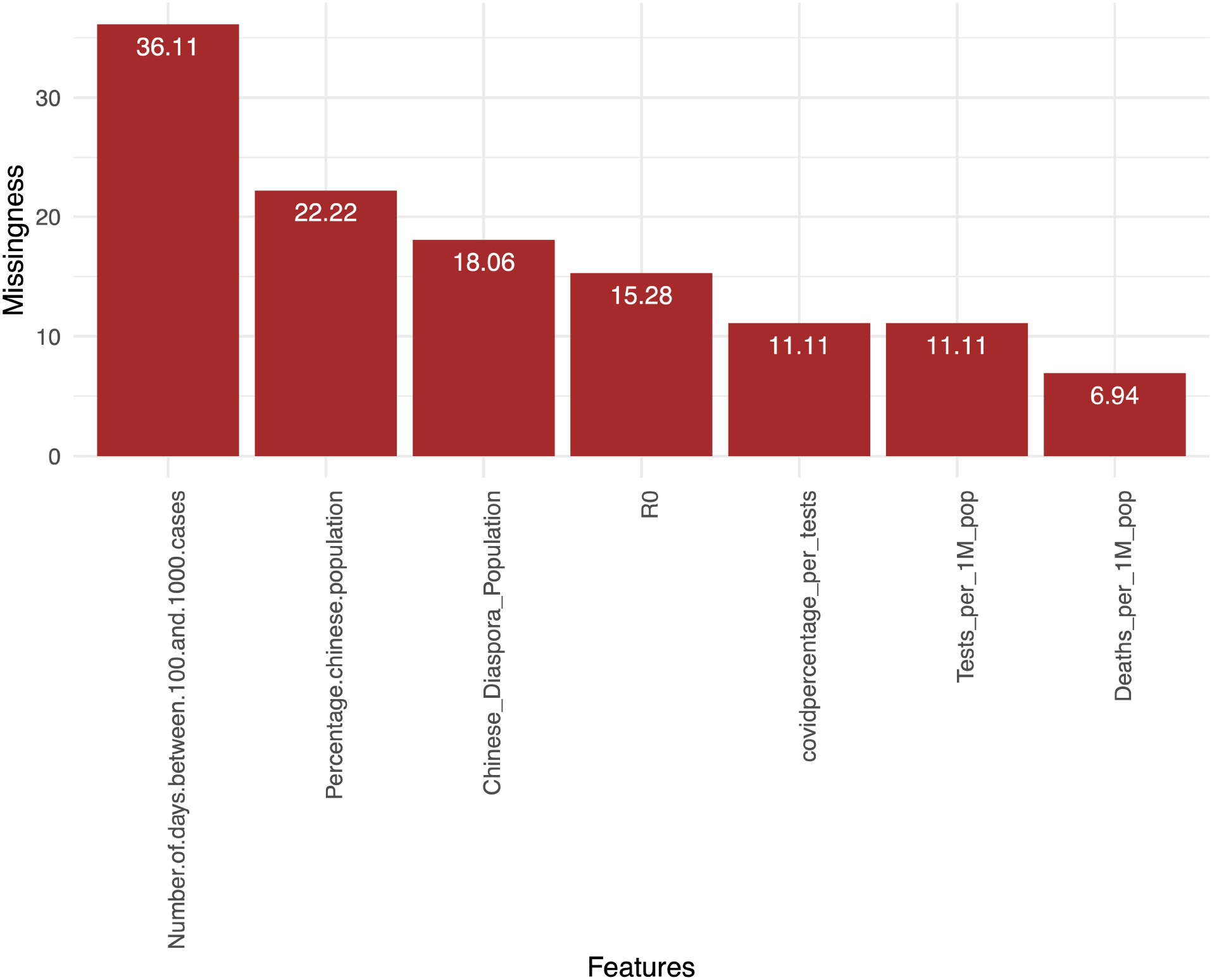
Missingness per variable. Since all the variables were within the acceptable band of missingness, a random forest based multivariate imputation algorithm was applied to impute the missing values in the data.

**Supplementary Figure 2:**
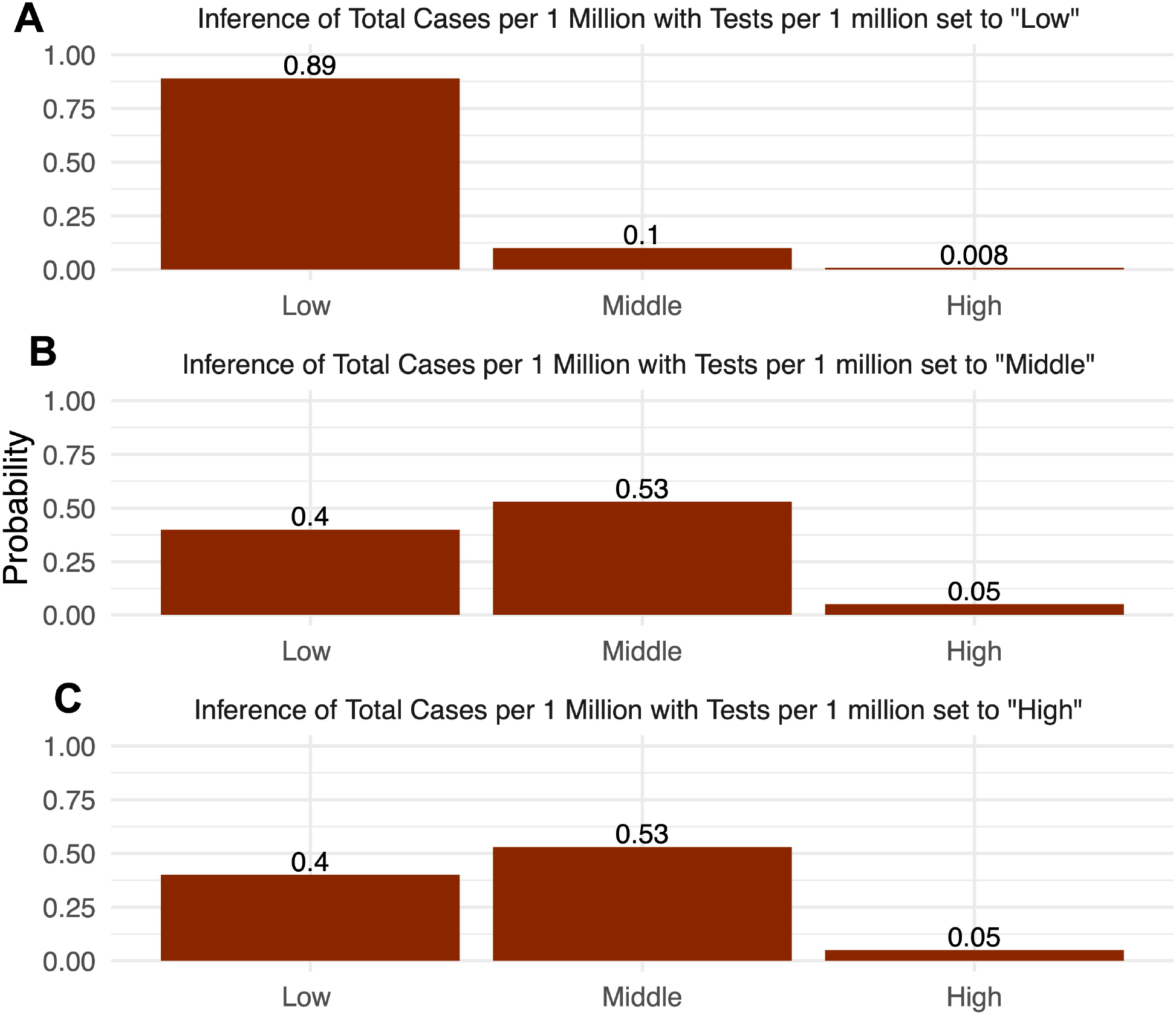
Positive non-monotonic correlation between testing rate and *Total cases per 1Milion Population*. Low testing -> low total burden; Middle and High Testing rate-> Nearly similar total number of cases.

## Notes

### Competing Interest Statement

The authors have declared no competing interest.

